# PLASMA H3.1-NUCLEOSOMES TO CLASSIFY SEVERITY AND SURROGATE RESPONSE TO TREATMENT IN HIDRADENITIS SUPPURATIVA: A COHORT STUDY

**DOI:** 10.64898/2026.01.13.26343988

**Authors:** Styliani Theohari, Aggeliki Vlyssidou, Anastasia Kourtesa, Argyro Anastasaki, Theodora Kanni, Pavlina Messiri, Evangelos J. Giamarellos-Bourboulis

**Affiliations:** 4th Department of Internal Medicine, National and Kapodistrian University of Athens, Medical School, Greece; Hellenic Institute for the Study of Sepsis, Athens, Greece

**Keywords:** neutrophil extracellular traps, Hidradenitis Suppurativa, HiSCR response

## Abstract

**Background:** Huge neutrophilic infiltrates within lesional and perilesional tissue in hidradenitis suppurativa (HS) give rise to the hypothesis that neutrophil extracellular trap (NET) formation may further drive systemic immune activation in HS. As intrinsic constituents of NETs, nucleosomes-particularly circulating nucleosome containing Histone H3.1 (H3.1-nucleosomes)-serve as reliable indicators of NETosis in the blood.

**Objectives:** To investigate whether plasma H3.1-nucleosomes, fluctuate with HS activity.

**Methods:** Participants were adults with moderate to severe HS. Peripheral blood samples were collected at each visit, EDTA plasma was prepared under standardized conditions and stored at -80°C. They were subsequently analyzed for circulating H3.1-nucleosomes with a proprietary assay. Baseline and longitudinal data were evaluated in relation to HS severity, clinical characteristics and clinical response using both the HS clinical response score (HiSCR) and the at least 55% decrease of the international HS4 score (IHS4-55).

**Results:** 93 patients were enrolled; in serial measurements were available for 54. Patients were classified into two clusters; hyper-H3.1 and hypo-H3.1 based on the over-time kinetics of H3.1-nucleosomes. The hyper-H3.1 cluster is characterized by more severe disease. H3.1-nucleosomes 24 ng/ml or more suggest higher total count of inflammatory lesions and of draining tunnels. More than 45% decrease of H3.1-nucleosomes between visits is associated with higher chances for attainment of HiSCR and IHS4-55 responses with biologicals targeting TNFα, IL-1 and IL-17.

**Conclusions:** Circulating H3.1-nucleosome levels reflect HS disease activity and surrogate response to treatment.

**What is already know about this topic?:** Biomarkers to provide precision approach in hidradenitis suppurativa remain an unmet need.

**What does this study add?:** For the first time an easy-to-measure blood test is presented to classify patients and to surrogate treatment. H3.1-nucleosomes distinguish patients into high- and low-level of neutrophil activation. Over-time decreases by 45% or more indicate response to biological treatment.

**What is the translational message?:** The new blood test may be used to initiate trials where treatment guidance of both initiation and early stop of treatment will be studied.

## INTRODUCTION

The introduction of biologicals was a major milestone not only in the management of hidradenitis suppurativa (HS) but also in the understanding of the pathogenesis. Clinical benefit coming from inhibition of tumor necrosis factor-alpha (TNFα) and of interleukin (IL)-17 shed light to the major role of both cytokines in HS pathogenesis. However, efficacy ranges between 40% and 60%^1–3^ and feeds several considerations. Among them questions on better understanding of HS pathogenesis and need of precision treatment start to emerge.^4^ Cytokine measurements in the pus indicate large variations between patients and give rise to the concept that not all patients fit the same model of management.^5^ This is aligned with the large over-time variations of the lesions of HS, which may appear even without treatment.^6^

HS is beyond any doubt a chronic neutrophilic disease. Neutrophils infiltrate around follicular structures, sinus tract formations and perilesional skin.^7, 8^ Neutrophils are the effector cell of NETosis, a cellular process involving chromatin recondensation, histone citrullination, rupture of the nuclear envelope and subsequent release of nuclear contents.^9^ Release of NETs is suggested to be detrimental in tissue inflammation and deformity and blood markers are myeloperoxidase (MPO)-DNA complexes, citrullinated histones (H3.1), neutrophil elastase (NE) DNA complexes and nucleosomes.^10, 11^ As intrinsic constituents of NETs, nucleosomes-particularly circulating nucleosome containing Histone H3.1 (H3.1-nucleosomes)-serve as reliable indicators of NETosis in the blood.^12^

We hypothesized that heterogeneity of treatment responses and large fluctuations of HS may associate with the activity of neutrophils for NETosis. As such, we measured NETs prospectively in a cohort of patients with moderate to severe HS and we associated responses to biological treatment with changes of H3.1-nucleosomes.

## PATIENTS AND METHODS

### Patient cohort

In this prospective cohort study, participants were under follow-up at the outpatient Department of Immunology of Infectious Diseases, of ATTIKON University General Hospital. The study was approved by the Ethics Committee of the hospital (approval 466/27-06-2023) and conducted according to the Declaration of Helsinki Principles. Written informed consent was obtained by all study participants.

Study participants were adults (age ≥18 years) of either sex with moderate to severe HS diagnosed for more than one year necessitating biological therapy (either adalimumab or secukinumab). Exclusion criteria were: a) any active viral, fungal or bacterial infection the last three months; b) chronic intake of corticosteroids defined as more than 0.4 mg/kg equivalent prednisone daily the last 30 days; c) neutropenia defined as absolute neutrophil count less than 1,000/mm^3^; d) any malignancy; and e) any other chronic inflammatory disorder including but not limited to systemic lupus erythematosus, rheumatoid arthritis and inflammatory bowel disease.

Blood was collection was performed during the first visit and repeated on each patient visit for one entire year. The time interval between visits was not standard for all patients. Patients were advised for follow-up visits every 3 months; they were also allowed for visits on demand. During each visit, patients were clinically evaluated, and a 4 mL blood sample was drawn from a peripheral forearm vein under aseptic conditions. Samples were collected into a 4 mL tube, coated with ethylenediaminetetraacetic acid (EDTA) and transferred to the laboratory within less than one hour from blood draw, under cool conditions (2-8^0^C). Blood was centrifuged at 400 x g for 10 minutes, at room temperature. Plasma was collected under aseptic conditions, taking care not to disturb the cell pellet and especially the white blood cell buffy coat; transferred to an Eppendorf LoBind® tube to ensure the reduced sample-to-surface binding and the maximal recovery of DNA and DNA-containing molecules. All samples were stored at -80°C and shipped to Belgian Volition SRL, (Isnes, Belgium) in dry ice, where they were analyzed for NETs concentration using a proprietary Nu.Q^®^ NETs assay, utilizing anti-histone H3.1 and conformational anti-nucleosome antibodies.

Recorded clinical data were demographics, age at HS onset, the total body count of inflammatory nodules (INs), abscesses and draining tunnels (dTs), Hurley stage and the type of treatment. The total body count of the inflammatory lesions was the sum of the total count of INs and of abscesses. Disease severity was expressed by the international HS severity (IHS4) score.^13^ The exact time interval between visits was provided in days.

### Study endpoints

The endpoints of the study were: a) the classification of patients with HS into clusters of NETosis based on the kinetics of H3.1-nucleosomes; b) the association of single H3.1-nucleosome measurements with HS lesions; and c) the association between over-time changes of blood H3.1-nucleosomes and HS clinical response. Clinical response was defined using both introduced measures of efficacy, namely the HS Clinical Response (HiSCR)^14^ and IHS4-55.^15,16^ HiSCR was defined as any at least 50% decrease of the total AN count without any increase of the total count of abscesses and of dTs. IHS4-55 was defined as any at least 55% decrease of the total IHS4 score from start of treatment. The time to the attainment of HiSCR and of IHS4-55 was also recorded in days.

### Statistical analysis

Variables with normal distribution were expressed as means (SD) and with non-normal distribution as medians and quartiles or as means (SE). In order to classify patients into clusters of high and low levels of NETosis, all H3.1-nucleosome values available from all patients for which serial measurements were available were analyzed. The median of the distribution of all values was calculated and patients were divided into the Hyper-H3.1 cluster when at least 30% of the available serial measurements was above the median; and into the Hypo-3.1 cluster when less than 30% of the available serial measurements was above the median. Patients with only two available serial measurements were classified into the Hyper-3.1 when both measurements were above the median. Patients with only three available serial measurements were classified into the Hyper-3.1 when two measurements were above the median. Comparisons of the maximum and minimum counts of ANs, dTs, IHS4 scores, scar counts, clinical response and white blood cells counts were done by the Mann-Whitney U test.

To associate H3.1-nucleosomes with HS lesions, we used data of all participants coming from their first visit. Patients were split into those with H3.1-nucleosomes above and below the median value calculated above. Comparisons between patients for the total counts of ANs and dTs were done by ordinal regression analysis; odds ratios (ORs) and 95% confidence intervals (CIs) were calculated. Making the hypothesis that 60% of patients with values above the median of H3.1-nucleosome would have inflammatory lesions and that 30% of patients with values above the median of H3.1-nucleosome would have inflammatory lesions, to demonstrate the difference with 5% power at the 90% level of significance, 92 patients should be enrolled in total.

In order to investigate if changes of H3.1-nucleosomes may surrogate response to treatment, the attainment or not of HiSCR and IHS4-55 was evaluated for patients with available serial measurements of H3.1. The percent difference between the maximum and minimum recorded H3.1-nucleosomes was calculated for every study participant. One receiver operator characteristics (ROC) curve was plotted for the prediction of HiSCR by the % calculated difference. The Youden index of the ROC was calculated as the cut-off with the best trade of sensitivity and specificity for prediction of HiSCR. Comparisons for the attainment of HiSCR were done for patients with % differences above and below the cut-off; ORs and 95%CIs were calculated by Mantel-Haenszel statistics. The time to attainment of HiSCR was compared by Cox regression analysis; hazard ration (HR) and 95% CIs were calculated.

Any value of p<0.05 was considered statistically significant.

## RESULTS

A total of 93 patients were enrolled between July 2023 and April 2025. Among these patients, serial measurements of H3.1-nucleosomes were available for 54 patients and only single measurements of H3.1-nucleosomes were available for 39 patients. No differences were found in HS severity and demographics between patients with serial and single measurements making plausible to extrapolate the analysis of the 54 patients to the entire cohort. In total 213 H3.1-nucleosome measurements were available for the 54 patients. The median of the 213 measurements was 24 ng/ml. Twenty-five were then classified into the Hyper-H3.1 cluster and 29 patients into the Hypo-H3.1 cluster using the classification criteria defined above. The two clusters presented significant differences. Both the maximum and minimum count of ANs, dTs and IHS4 were higher in the Hyper-H3.1 cluster. No differences between clusters were found for the total count of scars, the likelihood for the attainment of HiSCR and the maximum absolute count of neutrophils and monocytes (Figure 1). Mean (SE) years since HS onset were 7.86 (1.14) in the Hypo-H3.1 cluster and 7.92 (1.07) in the Hypo-H3.1 cluster (p: 0.971).

**Figure 1.**
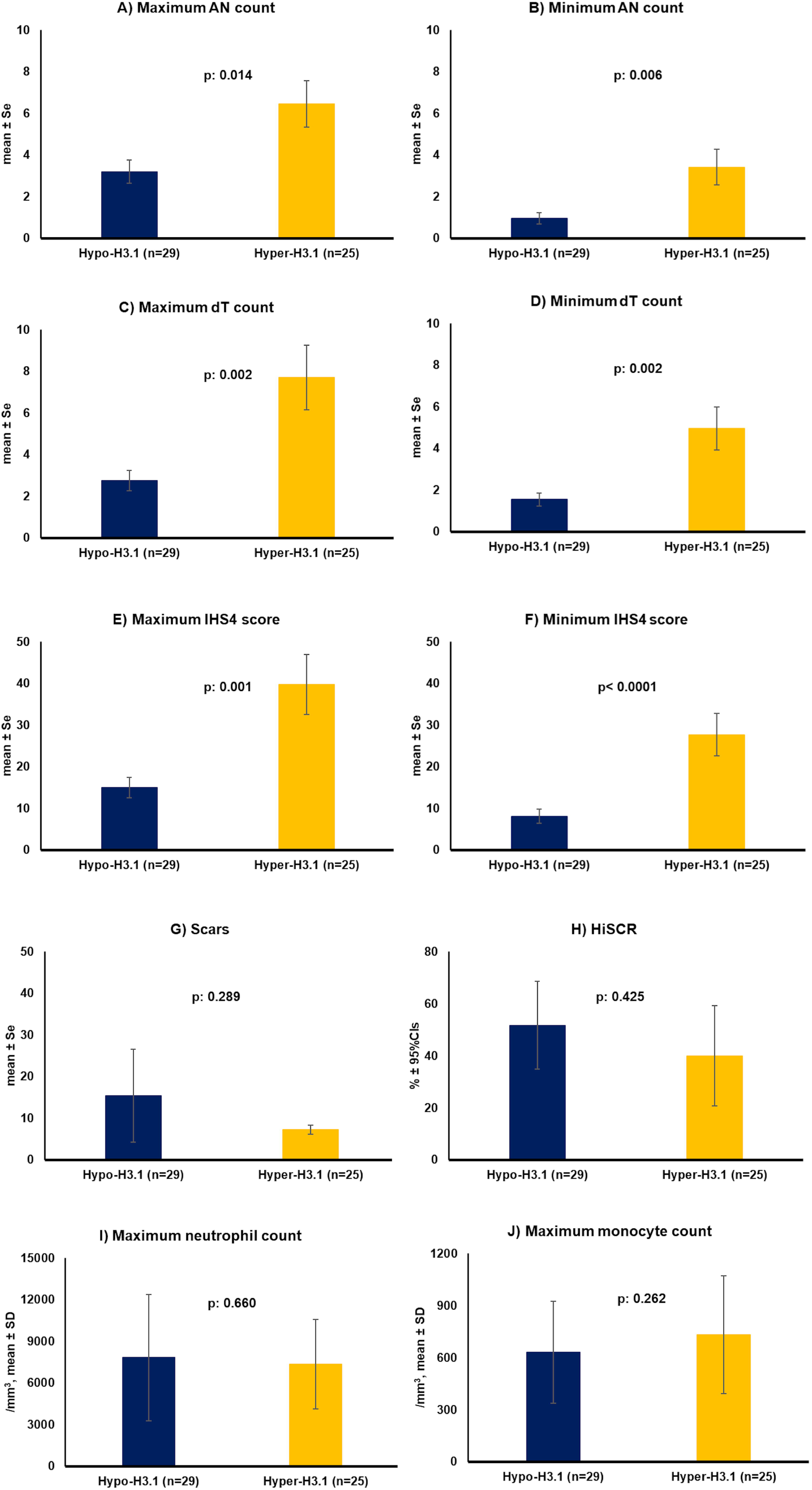
Characteristics of the hyper-H3.1 and hypo-H3.1 clusters of HS. A) Comparisons of the maximum over-follow-up AN counts between the two clusters B) Comparisons of the minimum over-follow-up AN counts between the two clusters C) Comparisons of the maximum over-follow-up dT counts between the two clusters D) Comparisons of the minimum over-follow-up dT counts between the two clusters E) Comparisons of the maximum over-follow-up IHS4 score between the two clusters F) Comparisons of the minimum over-follow-up IHS4 score between the two clusters G) Comparisons of the total count of scars between the two clusters H) Comparisons of the rate of patients attaining HiSCR between the two clusters I) Comparisons of the maximum neutrophil counts over-follow-up between the two clusters J) Comparisons of the maximum monocyte counts over-follow-up between the two clusters The p-values of comparisons are provided in each panel <u>Abbreviations</u> AN: total inflammatory lesion count (sum of inflammatory nodules and abscesses); CI: confidence interval; dT: draining tunnels; HS: hidradenitis suppurativa; HiSCR: HS clinical response score; IHS4: international HS4 score; n: number of patients; SD: standard deviation; SE: standard error

At a next step, we investigated if single measurements of H3.1-nucleosomes at concentrations ≥24 ng/ml indicate specific HS lesions. For this analysis, we used only the first measurement performed in every patient. Ordinal regression analysis showed that the presence of H3.1-nucleosomes at concentrations ≥24 ng/ml was associated with higher risk for increase of dTs and with borderline risk for increase of ANs (Figure 2).

**Figure 2.**
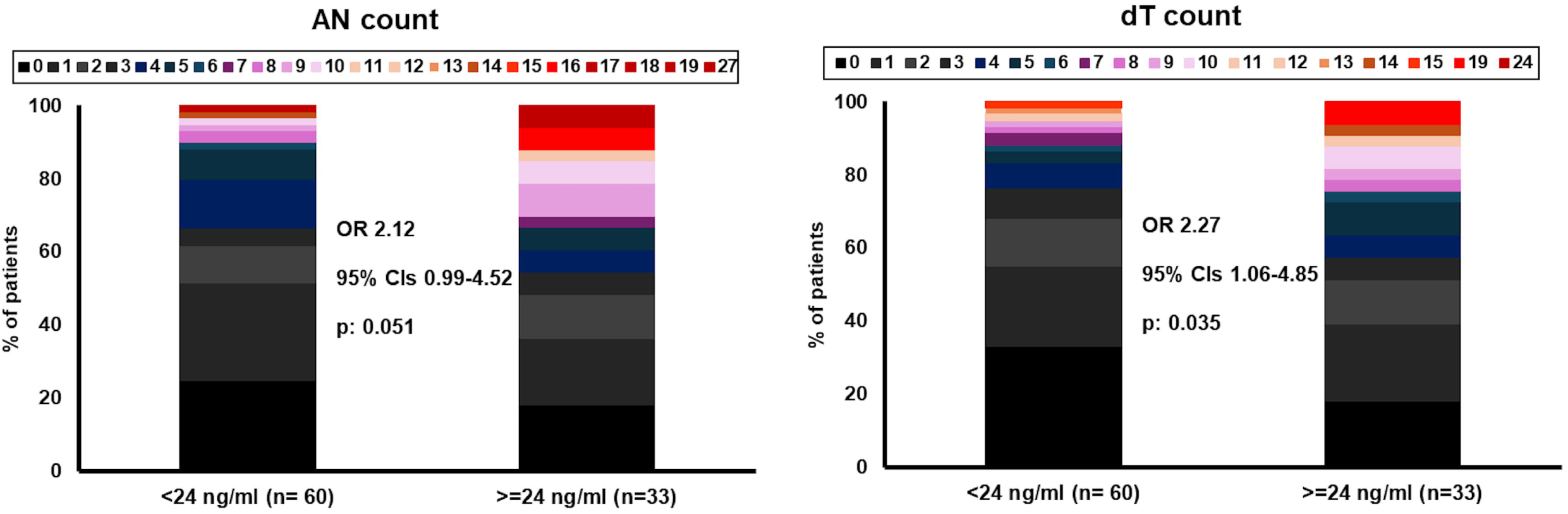
Counts of total inflammatory lesions (ANs) and of the draining tunnels (dTs) in relation to the H3.1-nucleosomes. Comparisons between patients with concentrations of H3.1-nucleosomes less than 24 ng/ml and ≥24 ng/ml are provided <u>Abbreviations</u> CI: confidence interval; OR: odds ratio

Using the optimal cutoff determined by Youden’s index, it was found that more than 45% decrease in H3.1-nucleosomes was predictive of the attainment of HiSCR. At this threshold, 61.8% of patients who experienced >45% decrease achieved HiSCR, compared to 20% of patients with <45% decrease (p = 0.004; Figure 3A). Furthermore, patients experiencing more than 45% reductions in H3.1-nucleosomes achieved HiSCR faster (Figure 3B). This level of decrease of H3.1-nucleosomes was attained with all three types of biologicals used for the treatment of study participants. The median time to reach >45% decrease in H3.1-nucleosome levels was 85 days in eventual HiSCR responders, compared to 155 days among non-HiSCR responders (Supplementary Figure 1). The time to decrease of H3.1-nucleosome concentrations was also faster among HiSCR responders (Supplementary Table 1).

**Figure 3.**
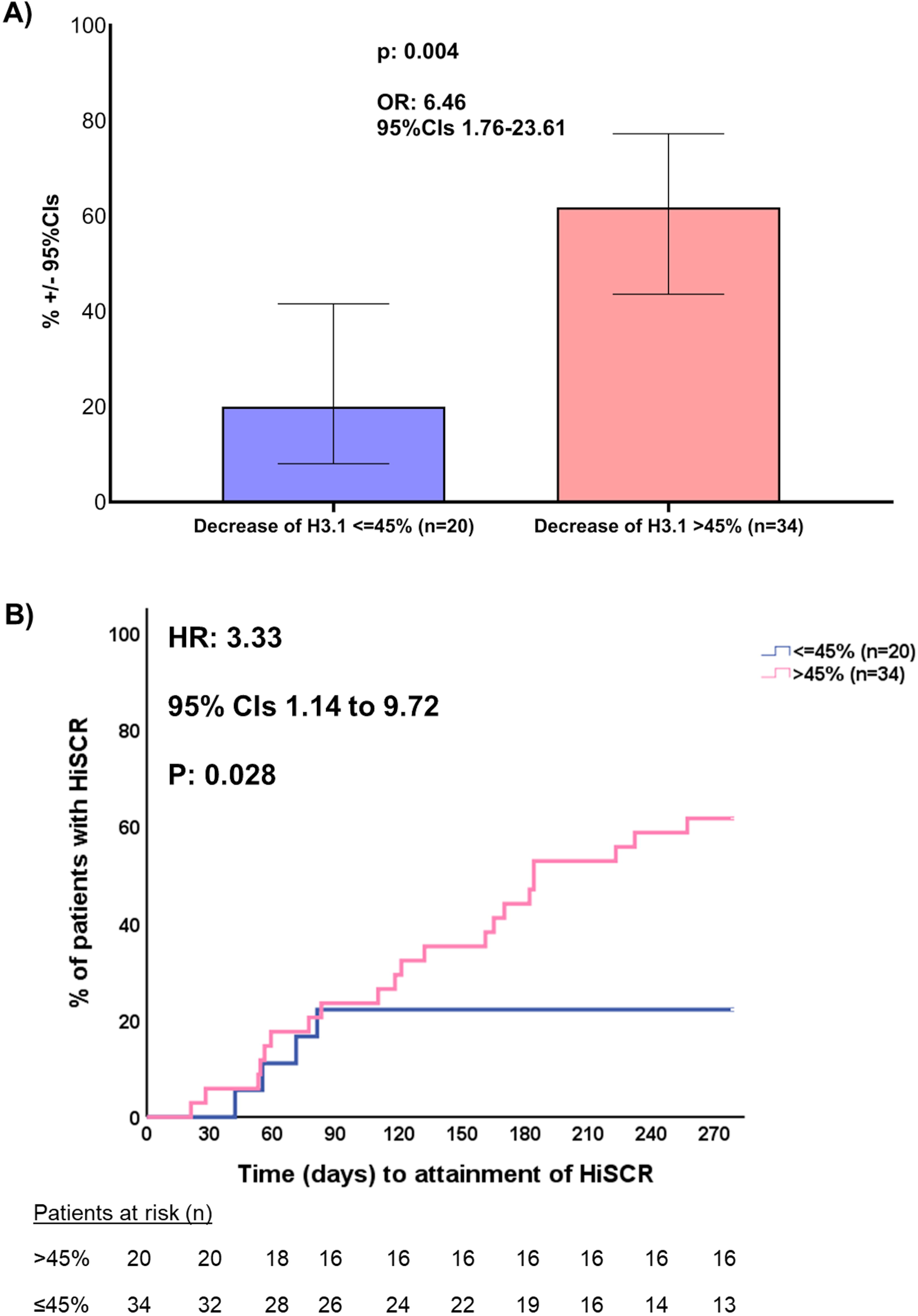
Association between decrease of H3.1 and attainment of HiSCR. A) % of HiSCR among patients experiencing more than 45% decrease of H3.1 and among patients experiencing ≤45% decrease of H3.1 B) Time to attainment of HiSCR among patients experiencing more than 45% decrease of H3.1 and among patients experiencing ≤45% decrease of H3.1 The p-values of comparisons between patients experiencing more than 45% decrease of H3.1 and among patients experiencing ≤45% decrease of H3.1 are provided in each panel <u>Abbreviations</u> CI: confidence interval; HS: hidradenitis suppurativa; HiSCR: HS clinical response score; HR: hazard ratio; OR: odds ratio; n: number of patients

Using the optimal cutoff determined by Youden’s index, it was found that more than 45% decrease in H3.1-nucleosomes was also predictive of the attainment of IHS4-55. At this threshold, 50% of patients who experienced >45% decrease achieved HiSCR, compared to 20% of patients with <45% decrease (p = 0.043; Figure 4).

**Figure 4.**
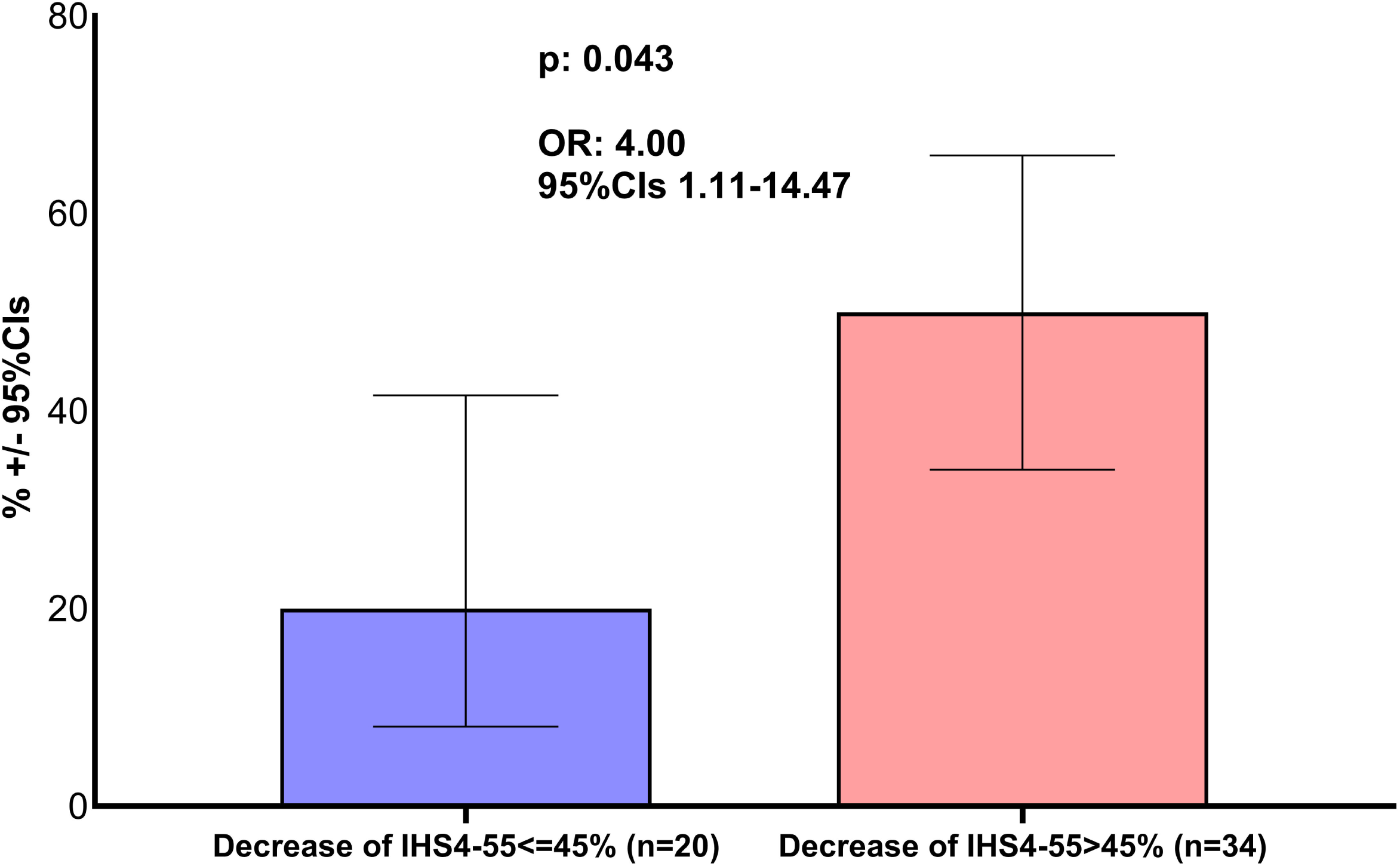
Association between decrease of H3.1-nucleosomes and attainment of IHS4-55. <u>Abbreviations</u> CI: confidence interval; HS: hidradenitis suppurativa; IHS4-55 decrease of international HS4 score 55% or more; OR: odds ratio; n: number of patients

**Table 1.** Demographics and disease characteristics of the 93 study participants. Patients are split into those with available serial measurements and into those with single measurements of H3.1

|  | **Patients with single measurements (N=39)** | **Patients with serial measurements (N=54)** | **p-value** |
| --- | --- | --- | --- |
| Age (years), mean (SD) | 40.0 (11.8) | 34.8 (12.2) | 0.053 |
| Age at HS onset (years), mean (SD) | 28.5 (12.5) | 26.9 (12.4) | 0.563 |
| Male/Female, n (%) | 19 (48.7)/20 (51.3) | 30 (55.6)/24 (44.4) | 0.535 |
| BMI, kg/m<sup>2</sup>, mean (SD) | 29.5 (7.3) | 30.1 (5.7) | 0.709 |
| Smoking, n (%) | 23 (59.0) | 38 (70.4%) | 0.276 |
| IHS4 score, mean (SE) | 17.5 (3.5) | 23.5 (3.6) | 0.255 |
| AN count, mean (SE) | 4.94 (1.09) | 3.85 (0.58) | 0.349 |
| Abscess count, mean (SD) | 2.17 (0.65) | 2.17 (0.30) | 0.994 |
| dT count, mean (SD) | 2.80 (0.54) | 6.62 (1.77) | 0.091 |
| Scar count, mean (SD) | 11.00 (2.75) | 20.84 (4.93) | 0.195 |
| Hurley score classification, n (%) |  |  |  |
| I | 2 (5.1) | 4 (7.1) | 1.00 |
| II | 26 (66.7) | 19 (35.2) | 0.003 |
| III | 11 (28.2) | 31 (57.4) | 0.006 |
| IHS4 classification, n (%) |  |  |  |
| Moderate | 17 (47.2) | 17 (34.0) | 0.266 |
| Severe | 19 (52.8) | 33 (66.0) |  |
| Absolute neutrophil count, /mm<sup>3</sup>,<br>mean (SD) | 7378.3 (3220.1) | 6504.2 (3192.1) | 0.250 |
| Absolute monocyte count, /mm<sup>3</sup>,<br>mean (SD) | 669.8 (231.7) | 597.9 (274.1) | 0.242 |

Abbreviations: AN, inflammatory lesions (sum of inflammatory nodules and abscesses); HS, hidradenitis suppurativa; BMI, body mass index; IHS4, international HS 4 score; n, number of patients; SD, standard deviation; SE, standard error

## DISCUSSION

HS is a heterogeneous skin disorder with systemic complications including major cardiovascular risk and other autoimmune phenomena like spondylodiskitis. This is reflected by the response rate to treatment with biological agents which block cytokines. The clinical efficacy of biological treatment never exceeds 60%^1–3^ whereas several times placebo responses as high as 47% are noticed.^17^ This paradox may come either from the diversity of immune activation between patients leading to large spontaneous fluctuations of lesions.^6^ One way to overcome this clinical obstacle is to use a biomarker. Biomarkers should be easily measurable, preferably in the blood, and they should make feasible to classify patients and surrogate response to treatment. No biomarkers with this characteristic have been described so far for HS.

In this longitudinal cohort of patients with moderate to severe HS, we demonstrated that H3.1-nucleosomes can help classify patients into two clusters based on their degree of NETosis: the hyper-H3.1 cluster and the hypo-H3.1 cluster which also reflect that most of the time course of the disease H3.1-nucleosome levels remain stable in the blood above or below 24 ng/ml respectively. The Hyper-H3.1 cluster is associated with higher AN and dT counts. Surprisingly the two clusters cannot predict the attainment of HiSCR with biological treatment. Instead over-time decreases can surrogate both HiSCR and IHS4-55 responses. Patients reaching more than 45% decrease of H3.1-nucleosomes attain faster clinical response. The decrease of H3.1-nucleosomes over the follow-up of patients contradicts previous findings from 18 patients that circulating neutralizing antibodies do not allow the degradation of NETs.^18^ However, this study by Oliveira et al. did not involve serial measurements and associations with treatment responses; they also analyzed NETosis as an expression of the DNAse 1 activity. Surprisingly, the absolute neutrophil and monocyte counts were similar between the hyper-H3.1 and the hypo-H3.1 cluster. Neutrophils are a major reservoir for NET production. The lack of difference in the neutrophil count between the two clusters suggests that it is the level of neutrophil activation which is different between the two clusters.

The current study has two limitations. The first limitation is that while Histone H3.1 is a widely accepted marker for NET formation, it may be influenced by other sources of cell death or inflammation. We tried to eliminate these by clinically assessing and omitting patients with clinical signs of topical or systemic inflammation. The second limitation is that the time intervals between visits was not stable for all patients and some patients underwent only one visit. However, the lack of difference in HS characteristics between patients with one measurement and serial measurements make plausible the generalization of results.

One of the major limitations in HS research is the need for standardization for the reporting of lesions between groups of investigators and between clinicians.^18^ Practice of medicine in other disease areas suggests that biomarkers is a way to tackle this problem. Our findings suggest that incorporating measurements of H3.1-nucleosomes into future clinical trials and treatment decision making may enhance precision in the immunotherapeutic management of HS and also limit the high placebo responses.

## Funding Information

The study was funded by the Hellenic Institute for the Study of Sepsis. Measurements of H3.1-nucleosomes were funded by Belgian Volition, SRL, Belgium

## Conflicts of Interest

EJ Giamarellos-Bourboulis reports honoraria from Abbott Products Operations, bioMérieux, Brahms GmbH, GSK, InflaRx GmbH, Sobi and Xbiotech Inc; independent educational grants from Abbott Products Operations, bioMérieux Inc, Johnson & Johnson, MSD, UCB, Swedish Orphan Biovitrum AB; and funding from the Horizon 2020 European Grants ImmunoSep and RISCinCOVID and the Horizon Health grants EPIC-CROWN-2, POINT and Homi-Lung (granted to the Hellenic Institute for the Study of Sepsis). The other authors do not have any conflict of interest to disclose.

## Ethical Approval

The study was approved by the Ethics Committee of ATTIKON University General hospital (approval 466/27-06-2023)

## Ethics statement

the participants in this manuscript have given written informed consent to the publication of their case details

## Data availability statement

The data that support the findings of this study are available from the corresponding author upon request.

## Author contribution

AV, ST and AK collected clinical data, generated the study database, drafted the manuscript, revised the manuscript for intellectual content and gave approval for submission.

AA collected biosamples and supervised processing, revised the manuscript for intellectual content and gave approval for submission.

PM analyzed the data, revised the manuscript for intellectual content and gave approval for submission.

EJGB analyzed the data, drafted the manuscript, revised the manuscript for intellectual content and gave approval for submission.

## Notes

### Competing Interest Statement

EJ Giamarellos-Bourboulis reports honoraria from Abbott Products Operations, bioMerieux, Brahms GmbH, GSK, InflaRx GmbH, Sobi and Xbiotech Inc; independent educational grants from Abbott Products Operations, bioMerieux Inc, Johnson & Johnson, MSD, UCB, Swedish Orphan Biovitrum AB; and funding from the Horizon 2020 European Grants ImmunoSep and RISCinCOVID and the Horizon Health grants EPIC-CROWN-2, POINT and Homi-Lung (granted to the Hellenic Institute for the Study of Sepsis). The other authors do not have any conflict of interest to disclose.

