## SUPPLEMENTARY FIGURE 1 for "PLASMA H3.1-NUCLEOSOMES TO CLASSIFY SEVERITY AND SURROGATE RESPONSE TO TREATMENT IN HIDRADENITIS SUPPURATIVA: A COHORT STUDY"

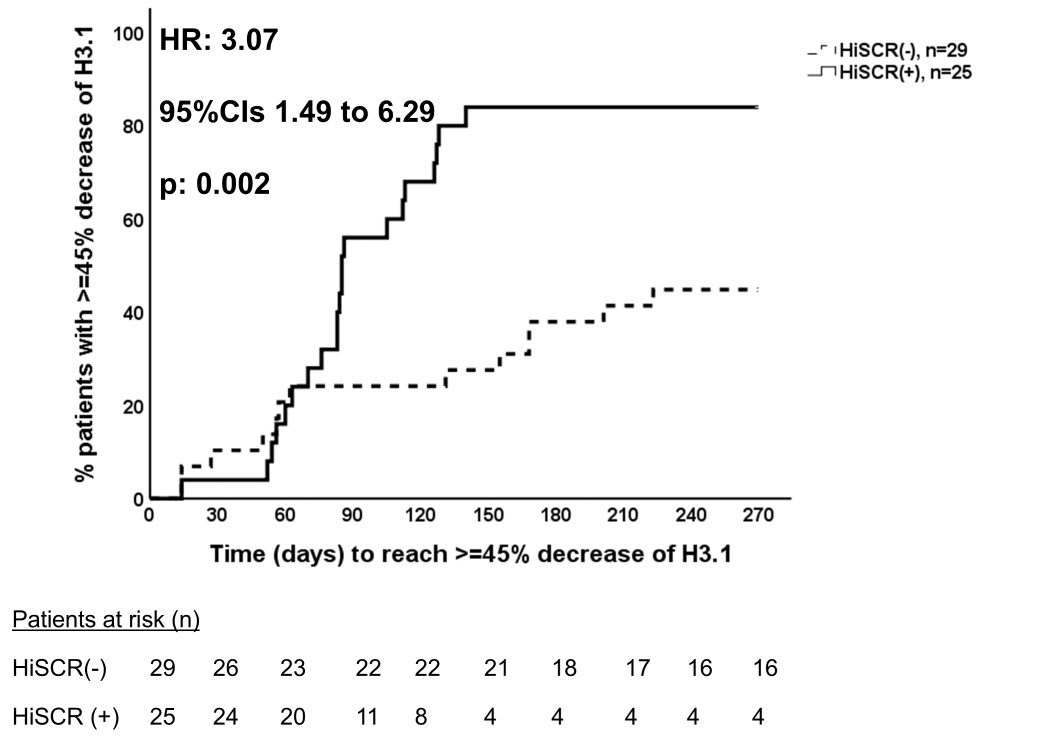

**Supplementary Figure 1 Time to decrease of H3.1 between patients who attained or not HiSCR**

The p-value of comparison between patients who attained HiSCR and patients who did not attain HiSCR is provided

Abbreviations CI: confidence interval; HS: hidradenitis suppurativa; HiSCR: HS clinical response score; HR: hazard ratio, n: number of patients
